# Adaptable Stroke Education Improves Knowledge Across Diverse High School Settings

**DOI:** 10.64898/2026.05.14.26353185

**Authors:** Samuel Namian, Rebecca DiBiase, Safa Hakim Elnazer, Chauncey Evers, Celia Fung, Rajiv Narula, Michael Rafferty, Adina Salahuddin, Daksh Jay Sardana, Jennifer Shea, Molly Sullivan, Rachel Forman

**Affiliations:** Department of Neurology, Yale School of Medicine, New Haven, CT, USA; Sevaro, New York, NY, USA; Derby Public Schools, Derby, CT, USA; Engineering and Science University Magnet School, West Haven, CT, USA; The Wheatley School, Old Westbury, NY, USA

## Abstract

**Background:** High school students may be able to communicate health topics to peers and adults. Yet, few studies have evaluated the role of high school students in community health initiatives, making them an underutilized group for disseminating health information. We pilot tested stroke education across five high schools using varied delivery approaches as a preliminary step toward evaluating youth stroke education to improve community health.

**Methods:** In April-May 2025, five high schools in Connecticut and New York participated in stroke education. The format was designed to fit the needs of each school and included an 8-session classroom curriculum (Derby, CT), after-school club meetings (New Haven, CT; Long Island, NY), and one large assembly (Bridgeport, CT). Developed by teachers and neurology providers, the curriculum covered stroke risk factors, symptoms, and emergency response. Students completed a 15-point assessment adapted from the validated Stroke Action Test before, immediately after, and 4-6 weeks post-intervention; data were collected between April and July 2025.

**Results:** Of 112 students completing the pre-test, 99 (88%) completed the immediate post-test and 51 (46%) the delayed follow-up. Average scores rose from 47% pre-intervention to 75% post and 70% at 4-6 weeks. All schools scored <50% on pre-tests suggesting poor baseline stroke knowledge.

**Conclusion:** This pilot suggests that stroke education can be delivered to high school students across varied settings and may support knowledge gains up to 6 weeks. Limitations included small sample sizes and missing follow-up data. If validated in larger studies, this adaptable, teacher-supported approach could offer a scalable public health strategy for improving community stroke preparedness.

## Background

Stroke is among the most time-sensitive clinical entities, yet approximately 70% of the 795,000 annual stroke patients in the United States arrive more than six hours after symptom onset, often rendering them ineligible for effective treatments.^1^ The most common reason for delay is failure to recognize symptoms and activate emergency services.^1^ This gap is more pronounced in socioeconomically disadvantaged communities, underscoring the need for targeted education.^2^

School-based programs such as Hip-Hop Stroke found that children in elementary school can learn to recognize stroke symptoms and transfer this knowledge to family members.^3^ Although these results are promising, high school students may offer even greater potential. Unlike younger children, they can engage with advanced content, share knowledge with both younger children and adults, and are beginning to make independent health decisions that shape their own stroke risk. Despite these potential advantages, high school students remain largely unstudied in stroke education research.

To address this gap, we pilot tested a stroke education curriculum^4^ across five high schools serving a diverse range of socioeconomic contexts. Rather than employing a single standardized format, we intentionally adapted our delivery approach to meet the logistical constraints and educational needs of each school setting, ranging from multi-session classroom curricula to single after-school sessions. This adaptive design allowed us to assess whether knowledge gains could be achieved across varied real-world implementation contexts. This pilot represents a preliminary step toward evaluating youth stroke education to improve community health.

## Methods

In April and May 2025, five high schools in Connecticut and New York participated in stroke education initiatives. The curriculum covered stroke pathophysiology, symptom recognition, modifiable risk factors, and emergency response, and included didactic and hands-on components.^4^ Delivery formats varied by school: Derby High School adapted the curriculum into an existing Anatomy and Physiology course that took place over 8 classes (Derby, CT); Hill Regional Career High School in New Haven, CT implemented a 3-session after-school club series, Harding High School (Bridgeport, CT) held a single large assembly for health students, and ESUMS (New Haven, CT) and Wheatley (Long Island, NY) had a single after-school science club session. Besides Derby High School, Yale clinicians taught the curriculum. Schools from varied socioeconomic contexts were selected to reach a racially and economically diverse student population, including students from communities disproportionately burdened by stroke. Students completed a 15-item assessment adapted from the validated Stroke Action Test^5^ before the intervention, immediately after, and 4-6 weeks after.

## Results

Of 112 students completing the pre-test, 99 (88%) completed the immediate post-test and 51 (46%) completed the delayed follow-up (Table 1). Mean scores increased from 47% at baseline to 75% immediately post-intervention and remained elevated at 70% at 4 to 6 weeks. All schools demonstrated poor baseline stroke knowledge. No school exceeded 50% on pre-tests. Knowledge gains were observed across all delivery formats, with improvements ranging from 19 to 32 percentage points immediately post-intervention. At delayed follow-up, mean scores declined by only 5 percentage points from post-intervention levels, remaining 23 percentage points above baseline across schools.

**Table 1.** Stroke Knowledge Assessment Scores by School and Delivery Format. Five high schools participated in stroke education delivered across varied formats. Racial demographics and free and reduced lunch (FRL) percentage reflect school-level demographics obtained from the National Center for Education Statistics (2024-2025). Major racial/ethnic groups shown; percentages may not sum to 100% due to students identifying as multiracial or other categories. Scores represent mean percent correct on a 15-item stroke knowledge assessment adapted from the Stroke Action Test.^5^ Pre-test was administered before the intervention, post-test immediately after, and delayed follow-up at 4-6 weeks. Number of respondents at each timepoint shown in parentheses. “Unknown” represents students whose follow-up response submission could not be linked to a specific school.

| School (Location) | Delivery Format | FRL% | Racial Demographics | Pre-Test | Post-Test | Delayed Follow-up |
| --- | --- | --- | --- | --- | --- | --- |
| Engineering Science University Magnet School (New Haven, CT) | Single club session | 49 | 29% Black<br>27% White<br>25% Hispanic<br>10.5% Asian | 47% (n=8) | 79% (n=6) | 87% (n=4) |
| Hill Regional Career High School (New Haven, CT) | 3-session classroom | 82 | 45% Hispanic<br>39% Black<br>9% White<br>5% Asian | 46% (n=22) | 65% (n=20) | 67% (n=3) |
| Warren Harding High School (Bridgeport, CT) | Single assembly | 89 | 59% Hispanic<br>33% Black<br>5% White<br>1% Asian | 48% (n=54) | 79% (n=51) | 68% (n=33) |
| Derby High School (Derby, CT) | 8-session curriculum | 54 | 45% Hispanic<br>24% White<br>23% Black<br>3% Asian | 47% (n=13) | 74% (n=13) | 69% (n=6) |
| Wheatley High School (Long Island, NY) | Single club session | 4 | 51% White<br>36% Asian<br>7% Hispanic<br>1% Black | 46% (n=15) | 78% (n=9) | 78% (n=3) |
| Unknown | — | — | — | — | — | 80% (n=2) |
| Total |  |  |  | 47% (n=112) | 75% (n=99) | 70% (n=51) |

## Discussion

These preliminary findings suggest that stroke education can be successfully delivered to high school students across varied settings and may support knowledge retention for at least 4 weeks. The low baseline scores across all schools reinforce the need for stroke education in this age group. Notably, knowledge gains were observed regardless of delivery format, from single assemblies to 8-session curricula, suggesting that schools may have flexibility in implementing stroke education based on available resources and scheduling constraints. The Derby curriculum was developed and delivered by teachers with minimal external support, and several schools including Derby have expressed interest in repeating the program annually, suggesting that this model may be sustainable beyond a research setting.

This study includes several limitations. The sample size was small, and high attrition at follow-up (54%) limits conclusions about knowledge retention. Without a control group, observed gains may reflect test-retest effects rather than true learning. Additionally, we did not assess whether students shared stroke knowledge with family members or whether education would improve real-world emergency response.

Despite these limitations, these preliminary findings suggest that adaptable stroke education may improve knowledge across diverse high school settings. Future research with larger samples and longer follow-up periods is needed to determine optimal delivery formats and evaluate effects on community stroke awareness and emergency response. Empowering high school students to recognize stroke symptoms, activate emergency services, and share information with their families and communities represents a promising and scalable public health strategy.

## Data Availability

All data produced in the present study are available upon reasonable request to the authors.

